# ROBUST COVID-19-RELATED CONDITION CLASSIFICATION NETWORK

**DOI:** 10.1101/2020.05.19.20106336

**Authors:** M Lucius, M Belvisi, CM Galmarini

**Author notes:** **Corresponding author:** Carlos M. Galmarini MD, PhD, Topazium Artificial Intelligence, P. de la Castellana 40 PI8, 28046 Madrid, Spain.

## Abstract

COVID-19 can exponentially precipitate life-threatening emergencies as witnessed during the recent spreading of a novel coronavirus infection which can rapidly evolve into lung collapse and respiratory distress (among other various severe clinical conditions). Our study evaluates the performance of a tailor-designed deep convolutional network on the tasks of early detection and localization of radiological signs associated to COVID-19 on frontal chest X-rays. We also asses the framework’s capacity in differentiating the above-mentioned signs, which are usually confused with the more usual common bacterial and viral pneumonias. Open-source chest X-ray images categorized as Normal, Non-COVID-19 and COVID-19 pneumonias were downloaded from the NIH (n=2,259), RSNA (n=600) and HM Hospitales (n=2,307). Our algorithmic framework was able to precisely detect the images with COVID19-related radiological findings (mean Accuracy: 90.5%; Sensitivity: 80.6%; Specificity: 98.0%), whilst correctly categorizing images deemed as Non-COVID-19 pneumonias (mean Accuracy: 88.4%; Sensitivity: 93.3%; Specificity: 92.0%) and normal chest X-rays (mean Accuracy 92.1%; Sensitivity: 91.8%; Specificity: 94.3%). The associated results show that our AI framework is able to classify COVID-19 accurately, making of it a potential tool to improve the diagnostic performance across primary-care centres and, to grant priority to a subset of algorithmic selected images for urgent follow-on expert review. This would sensibly accelerate diagnosis in remote locations, reduce the bottleneck on specialized centres, and/or help to alleviate the needs on situations of scarcity in the availability of molecular tests.

## INTRODUCTION

Reverse-transcription polymerase chain reaction (rt-PCR) tests remain the gold-standard for COVID-19 diagnosis in the acute phase of infection ^1, 2^. However, this molecular diagnostic tool has some limitations: (i) its false negative rate is relatively high (20-60%); (ii) obtention of results varies from 4-6 hours to, sometimes, a few days; (iii) it requires to be performed by certified laboratories with trained personnel, expensive equipment and availability of reagents; (iv) a large demand can easily overcome supply, particularly at times when its needed the most ^3, 4^. The situation is even more complicated in remote areas affected by the pandemic or in those with access only to low complexity healthcare centres. These, generally, have difficulties in accessing the rt-PCR tests forcing non-specialized practitioners to diagnose the COVID-19 condition based solely on clinical and/or radiological data. Drawbacks like these, have lately driven the debate on using chest imaging as a primary diagnostic tool. Indeed, two studies have reported high sensitivity of non-contrast chest CT compared to rt-PCR ^5, 6^. However, because of the associated operational costs and limited equipment availability, chest CT can hardly be relied upon for initial diagnosis in mass. As a consequence, most clinical guidelines only recommend it when an alternative diagnosis is paramount and/or when COVID-19 testing kits are scarcely available. Similar attempts were recently performed, all based on chest x-rays which are widely available and less expensive to perform than CT equivalents. However, chest x-rays taken in patients with confirmed and symptomatic COVID-19 condition can induce to confusion in cases associated to other lung infections or pathologies (including the absence of them) making it difficult for non-trained physicians to differentiate among these patterns. Even in the case of expert radiologists, it has proven challenging to be precise on diagnosis (particularly with early stage patients).

The described scenario is what motivated us to design, code and optimize a robust COVID-19 classification network as a complementary tool to the current professional and equipment available resources.

Artificial intelligence (AI) tools aim to reproduce human cognition and processes involved in the analysis of complex data ^7^. Their use in assisting physicians has recently made inroads in various medical fields. Specifically, in the context of radiology, image recognition using a set of deep neural networks (“DNNs”) has proven to be a valuable aid to physicians diagnosing among many different pathologies. In radiology, these complex algorithms achieve accuracy rates comparable (or, in some instances, beyond) to those related to the ones by radiologists in most of the specific fields ^8^. Thus, AI-based analytical tools help securing increased precision along the entire spectrum of diagnostic radiology and, improving workflow prioritization in the case of large-scale screenings. Since the beginning of the current pandemic a few studies have been published proving the use of AI systems in diagnosing COVID-19 condition using radiological images ^9-13^. In differentiation to the existing research on the field, ours focuses on achieving improved measures of generalisation and stability across the different type of images provided by varied sources and equipment through a series of innovative algorithmic design features (details of which are beyond the scope of this work). These features also facilitate the accurate classification when images to be used for interpretation are gathered using an ordinary mobile phone camera exposed to chest X-rays projected by a negatoscope.

## MATERIAL AND METHODS

### Chest X-ray datasets

Our study is based on an aggregated pool of images sourced from HM Hospitales (n=2,307), the National Institute of Health (“NIH”; n=2,259) and the Radiological Society of North America (“RSNA”; n=600). In all cases, the files include anonymous frontal chest X-rays, whilst the dataset provided by HM Hospitales contains anonymized records related to the 2,307 patients admitted with a confirmed (n=2,075) or pending (n=232) of COVID-19 diagnosis performed by rt-PCR. All relevant ethical guidelines have been followed for the use of this material. The NIH dataset was downloaded from https://nihcc.app.box.com/v7ChestXray-NIHCC and the RSNA dataset was downloaded from https://www.rsna.org/en/education/ai-resources-and-training/ai-image-challenge/RSNA-Pneumonia-Detection-Challenge-2018 ^14^, ^15^. In the case of the images sourced from both the NIH and RSNA databases, only a subset of records was randomly selected. Figure 1 presents an example image for each of the labels found on the master dataset. Once aggregated to the files obtained from HM Hospitales, the resulting pool (n=5,166) was split into training (n=3,472), validation (n=1,051) and test (n=643) sets in a completely disjoint manner. In all instances, images contained across every bucket included a representative collection of the three classification categories: Normal, Non-COVID (including bacterial and other viral pneumonias) and COVID-19 pneumonias. The final composition for each dataset is detailed in Table 1.

**Figure 1.**
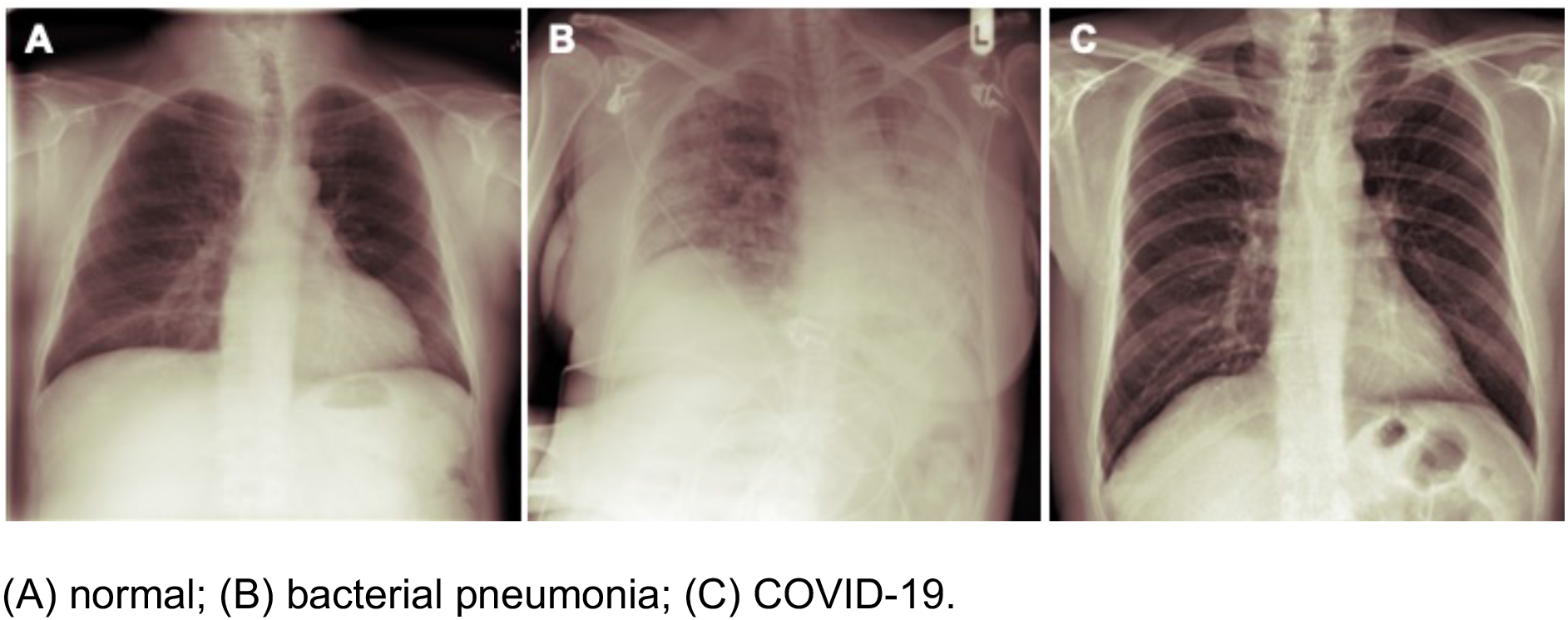
Examples of chest X-ray images from the master dataset.

**Table 1.**
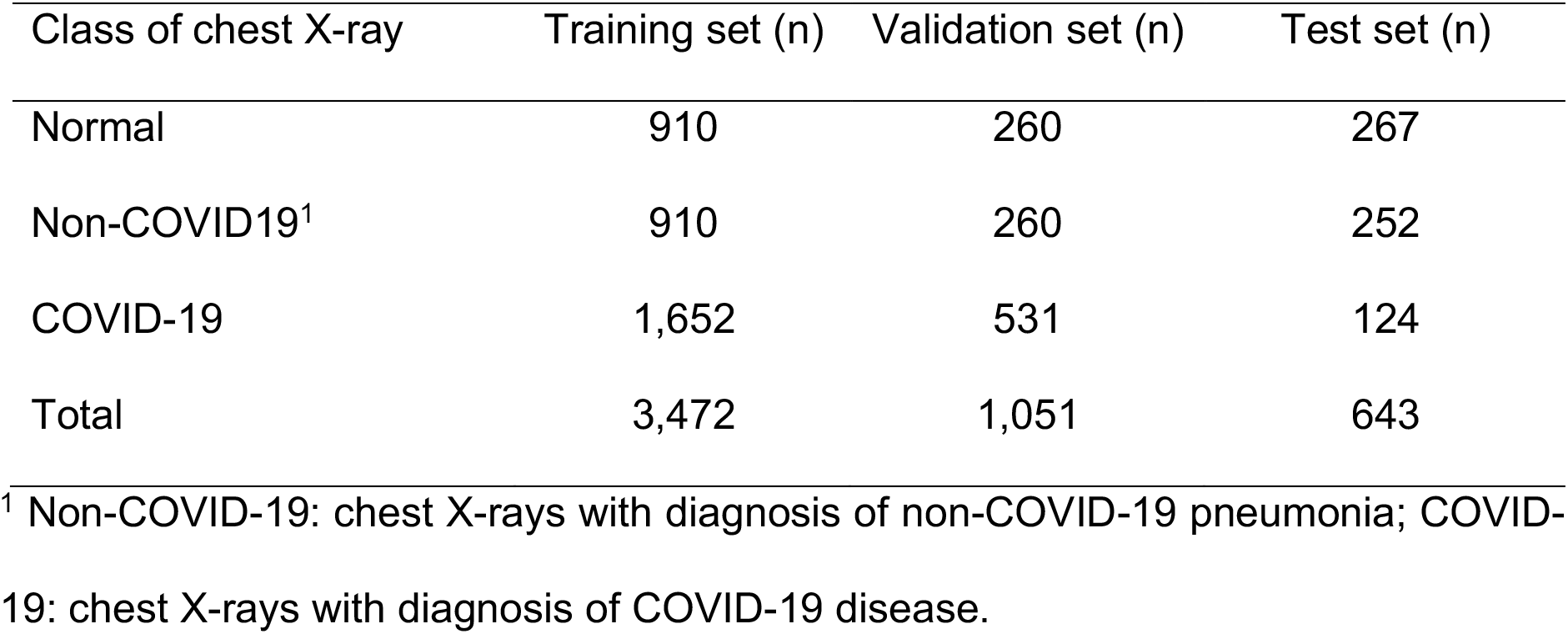
Description of the training, validation and test datasets.

### Network architecture

A Dense Convolutional Network architecture (“DenseNet”) has been the one accomplishing the most out of a comprehensive architectural network search initially performed. Such a design is known for delivering significant performance improvements over most of the alternative constructions, whilst requiring less memory and computational support. As described in previous publications, the DenseNet configuration is one where each layer is connected to every other layer through a feedforward linkage ^16^. At every level, feature-maps from all preceding layers are used as inputs, whilst its output feeds all subsequent deeper layers. The framework is composed by 4 dense blocks of 6, 12, and 24 layers each. In order to improve the performance and to optimize training, we have run calibration routines a total of 5 times (folds), each consisting on 200 epochs. Overall, training using the curated image patches took 55 h and 156k iterations to complete, using a 4 GeForce GTX 1080 GPU configuration. Maximum accuracy (100%) was reached after 125 epochs, whereas the pretrained version took 32 epochs for convergence to begin. The network was trained, validated and tested using images adjusted to 1024×1024×1 pixels/channel.

### Image pre-processing

When a deep convolutional neural network overfits, it performs extremely well on a training set but poorly on data outside of the calibration spectrum. In our design, two steps were taken to address this issue: (i) a dropout layer was added (set to 0.25) which results in 25% of the neurons to be randomly turned off during the training process, therefore reducing the likelihood of overfitting; (ii) a data augmentation process was performed where randomly selected data points were duplicated and modified to account for the variability found across different image capturing methods. To address variations related to grid location, size of the radiological finding and the image angle, all of the training images were altered using a combination of random rotation, zoom, shear, and flipping. The model was calibrated initially once just using dropout and, in a second occasion, both with data augmentation and dropout activated.

### Statistical analysis

Following the Training stage, a Validation process was performed in which images across the three different chest X-ray’s categories were used. Finally, a Test phase was run, which results were statistically analysed. A Confusion Matrix was constructed comparing the framework’s Classification Accuracy across each of the labels. Additionally, Accuracy, Error rate, Sensitivity, Specificity and Geometric Mean were estimated for each class ^17^. Accuracy defines the ratio between the correctly classified test samples (including each of the three labels) to the total number of test items ^17^. Error Rate was derived representing the complement of Accuracy. Sensitivity or True Positive Rate (“TPR”) represents correctly classified samples in relation to the total number of positive samples; Specificity or True Negative Rate (“TNR”) estimates the ratio of correctly classified negative samples with respect to the total universe within the same class. Geometric Mean (GM) was calculated by using the square root of the product of both TPR and TNR’s metrics. All these metrics are suitable standards to evaluate the classification performance on imbalanced data pools as the case is on this dataset. Finally, the above-mentioned estimates were calculated across each of the class labels documented in the corresponding original database archives.

## Results

### Patient population

We have had access to two basics demographic descriptors, gender and age, which were included on the original datasets. In the case of Normal X-rays, the data distribution related 43.7% to women and a 56.3% to men. The Non-COVID-19 split corresponded 40.6% to women and 59.4% to men, and in the case of COVID-19, 40.3% and 59.7% to women and men respectively. In terms of age distribution, the mean/SD estimated (expressed in years) was 46.7+16.4, 45.0+17.2 and 67.8+16.0 for each of the corresponding classification classes. As expected, and due to the COVID-19 suspected condition, the dataset originated on hospitalized patients shows a higher mean age when compared to the other two datasets.

### Classification metrics

Figure 2 presents examples of decision visualization on chest X-rays. Global Accuracy, Error Rate, TPR, TNR and GM for the predicted classification pool across all of the three categories are shown in Table 2. The overall Accuracy performance for the multi-classification task reached 90.4%. Our system was able to differentiate COVID-19 positive related chest X-ray images from the other two classes (common pneumonia and normal) with Accuracy, TPR and TNR of 90%, 80% and 98%, respectively. Of the 23 false negative patients, 9 were wrongly classified as Normal chest X-rays but, it is not clear whether these errors were a true misclassification or related to the fact that approximately 40% of COVID-19 positive tested patients are asymptomatic and do not usually show any observable radiological sign distinguishable by traditional screening methods. The remaining 14 cases were wrongly categorized as Non-COVID-19 pneumonia. In any case and, despite the minimal error in classification observed, when/if radiological signs of pneumonia are detected, a validation diagnosis should be performed by means of additional alternative tests. Finally, out of the 10 false positive cases, 5 were related to Normal chest X-rays and 5 corresponded to Non-COVID-19 pneumonia. For the Normal and Non-COVID19 classes, Accuracy rate reached 92% and 88%, respectively. In the case of Normal chest X-rays, estimated TPR and TNR were of 91% and 94%. As for the Non-COVID19 pneumonia instances, calculated TPR and TNR measurements were 93% and 92%.

**Figure 2.**
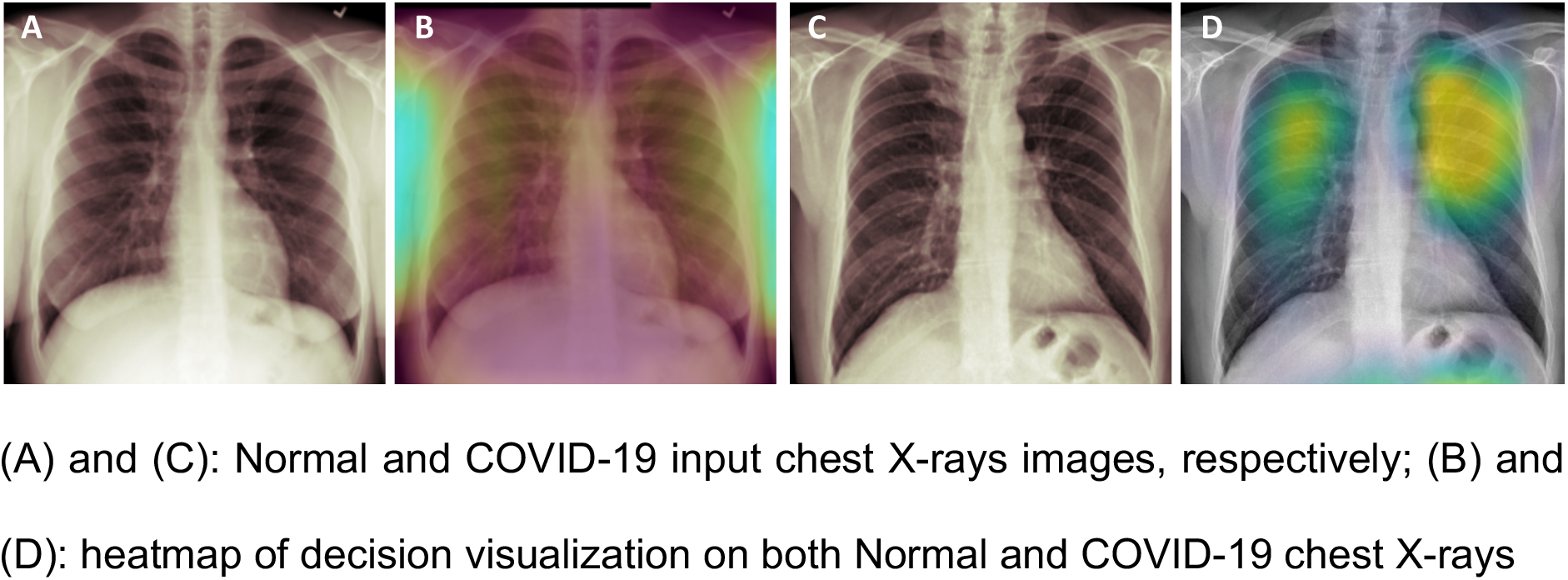
Examples of finding localization in radiographs using heatmaps.

**Table 2.** Classification metrics.

| DNN | Accuracy | Error rate | TPR<sup>1</sup> | TNR | GM |
| --- | --- | --- | --- | --- | --- |
| Normal | 0.921 | 0.078 | 0.918 | 0.943 | 0.930 |
| Non-COVID19<sup>2</sup> | 0.884 | 0.115 | 0.933 | 0.920 | 0.926 |
| COVID-19 | 0.905 | 0.094 | 0.806 | 0.980 | 0.889 |

<sup>1</sup> TPR: true positive rate; TNR: true negative rate; GM: geometric mean
<sup>2</sup> Non-COVID-19: chest X-rays with diagnosis of non-COVID-19 pneumonia; COVID-19: chest X-rays with diagnosis of COVID-19 disease.

## Discussion

Our results demonstrate that a deep learning framework trained on a diverse set of images can achieve classification Accuracy Rates in line (or beyond) the ones by specialized radiologists. The robust multi-class classification system presented has proven to be accurate across each and all of the three main classes (Normal, Non-COVID-19 and COVID-19). The use of AI applications as a diagnostic aiding tool is a growing trend in radiology providing a significative help among specialized professionals as well as general practitioners inducing the shortening of diagnostic workflow time and, an increase in diagnosis precision. These tools can also ease the exponential demand for diagnostic expertise during pandemic periods, letting radiologists to focus on the most urgent flow whilst allowing suspected patients within remote areas to get an immediate preliminary diagnosis. In addition, our system is designed to be able to process images obtained by standard mobile devices and nonprofessional cameras on chest X-rays projected on the negatoscope. Additionally, the framework could be also integrated into any existing teleradiology platform.

The advent of machine learning algorithms has made automated classification of radiological signs on chest X-rays an achievable target milestone. For example, CheXNet is a 121-layer CNN trained with ChestX-ray14, a large publicly available chest X-ray dataset containing over 110,000 frontal view X-ray images from 14 different diseases ^8^. Not only did CheXNet performed better than radiologists at diagnosing pneumonia, but it also overperformed when identifying other 13 diseases including cancer, pleura thickening and tuberculosis ^8^. Different machine learning systems have demonstrated their potential for diagnosing pediatric pneumonia using chest X-rays as well ^18^. In the case of COVID-19, previous studies demonstrated that deep learning architectures were able to detect positive cases accurately. Wang et al. showed that COVID-Net, an open-source deep convolutional neural network, achieved 92.6% accuracy in classifying normal, bacterial pneumonia, non-COVID viral pneumonia and COVID-19 viral pneumonia on 13,725 frontal chest radiographs. This study though included just 183 images out of 129 COVID-19 positive patients (COVIDx dataset) ^13^. Abbas et al. have used the DeTraC system achieving a 95% accuracy on a dataset of 196 chest X-rays including 105 patients COVID-19 positive patients ^9^. Using a pre-trained ResNet-50, Bukhari et al. demonstrated a 98% accuracy on a 278-image dataset including 89 COVID-19 positive patients whilst, on a modified version of the COVIDx dataset including 259 COVID-19 positive images, Karim et al reported 94% and 88% accuracy rates for DenseNet-161 balanced and imbalanced datasets, respectively ^11, 12^. Our results showing a 93% accuracy with a DenseNet on a dataset including 2,307 COVID-19 positive images are in line with the described antecedents, with the additional assurances provided by the disproportionally higher number of positive cases.

Also, based on the above, it is clear that an AI-based tool like ours can be of particular utility during stress and pandemic periods. This relies on the fact that COVID-19 condition exhibits particular radiological signatures and image patterns which can be observed in medical imagery and used for differential diagnosis from other pathologies ^19, 20^. Additionally, chest X-rays are the most widely utilised imaging method to establish the diagnosis of patients with respiratory problems, especially in rural and isolated areas with limited access to molecular tests or advanced medical equipment (such as CT scanners). Finally, in addition to the difficulties described, the demand overflow and workload pressure on expert radiologists usually witnessed during pandemic periods could potentially induce to perceptual and cognitive biases, all of which leads to random diagnostic errors ^21^. Besides, in hospitals and general healthcare centres lacking resident specialized radiologists, AI systems could prove useful in delivering an initial real-time diagnosis in any suspected patient just relying on X-ray equipment, a mobile application and access to a tele-radiology platform^22^.

Our framework was bound to only three different classes, which does not reflect the clinical reality of the many more conditions to be taken into account when diagnosing respiratory diseases on a chest X-ray. As a consequence, the use of our classification system should be regarded as an assisting tool for radiologists and/or general practitioners, aiming at improving accuracy within a limited context but, not as a replacement of qualified physicians diagnosis (when and if available). On the other hand, deep learning models are powerful “black box” models which remain relatively uninterpretable compared to the statistical kind traditionally used in medical practice. Computer vision models combine pixel-based visual information in a highly intricated way, making it difficult to trace the model output back to the observable input. In this sense, the use of heat maps highlighting critical regions on the chest (used as class-discriminating areas within the lungs) serve as a reassuring measure to both non-expert physicians and practising radiologists. Another limitation is the fact that although the test dataset was disjunct from the training dataset, all the COVID-19 positive images belonged to the same original database (HM Hospitales), raising concerns about the framework ability to generalize on exogenous test sets coming from different image banks. It is known that the efficacy of DNNs varies based on the set of images with which they are trained. Each model may have different sensitivities and specificities and may be subject to a unique set of biases and shortcomings in prediction introduced by the image training set. Finally, not all COVID-19 cases are associated with chest pathology. In fact, approximately half of patients imaged 0-2 days after symptom onset had a normal chest CT ^10^. We intend to address these limitations in future editions of this work.

## Conclusions

In summary, the results exposed show that AI-based tools can be used to discriminate between COVID-19 positive patients and the ones with other (or no) pulmonary infections. The system is designed to equally process images obtained by photographing X-rays using mobile devices (cell phones or tablets) or, scanning chest radiographs. Additionally, the framework could be integrated into tele-radiology systems via an automated-programming interface (API). In any case, the presented technology probes to be of extreme help in the context of accelerating the initial and preliminary detection of COVID-19-related pulmonary conditions when traditional molecular methods are temporally or permanently unavailable. Furthermore, the system can be adapted to assess the future evolution of the disease, that is to estimate probability that any given patient will aggravate from an initial diagnosed clinical condition potentially demanding a future admittance into specialized care units. Though a secondary feature, this could substantially ease the pressure on existing limited healthcare infrastructure by allowing practitioners to objectively filter through (for admittance) only patients whose condition is predicted to worsen. Ultimately, systems based on DNNs will allow general practitioners across undeserved and/or demand overflown regions, to act as a first point of diagnosis with expected accuracy rates similar, or in some instances better, to those achieved by specialized professionals and/or by means of utilising dedicated test kits.

## Data Availability

In this work, we have use from the public datasets from HM Hospitales, NIH and RSNA. The former dataset should be requested to HM Hospitales. The NIH and RSNA can be downloaded from the links provided in the Ms.

https://nihcc.app.box.com/v/ChestXray-NIHCC

https://www.rsna.org/en/education/ai-resources-and-training/ai-image-challenge/RSNA-Pneumonia-Detection-Challenge-2018

## Acknowledgements

We acknowledge the NIH Clinical Center, the RSNA and HM Hospitales for the access to their image datasets.

